# Development and Validation of MetaVidence: A No-Code Application for Accessible and Reproducible Meta-analysis

**DOI:** 10.64898/2026.11.17.26368328

**Authors:** Valbert Oliveira Costa Filho, Pedro Robson Costa Passos, Mariana Macambira Noronha, Rafael Lara Nohmi, Luis Felipe Leite da Silva, Erick F. Saldanha, Giuseppe G. F. Leite

## Abstract

**BACKGROUND:** Meta-analyses are central to evidence-based medicine, but the R packages that implement current methods require programming skills, and most graphical tools cover only a single study design. Here, we aimed to develop and validate a free no-code application that performs the main forms of meta-analysis within a single interface.

**METHODS:** We developed MetaVidence as a Shiny application built on the meta, metafor, netmeta and lme4 packages, which runs in the browser through WebAssembly and is also available as an R package. Agreement with direct package calls was evaluated across effect measures, models, estimators and secondary analyses, and 12 published meta-analyses were reconstructed. Usability was assessed with the System Usability Scale among researchers who had previously conducted a meta-analysis.

**RESULTS:** MetaVidence performs pairwise, single-arm, pre-calculated, network and diagnostic test accuracy meta-analysis through a three-step workflow of data verification, model specification and results. In three of the five families, multiple outcomes can be analyzed in a single run with the same settings, and every analysis reports the parameters that should be described in the methods section. A total of 6,321 validation checks were performed, and the maximum absolute difference in the reference-dataset comparisons was 3.9 × 10^−16^. The diagnostic module reproduced the anti-CCP example of the Cochrane Handbook to within 0.0002, and all 12 published meta-analyses were reproduced within half of the last reported digit. Fifty-nine researchers rated MetaVidence with a mean System Usability Scale score of 82.8 (95% CI 76.2-89.5).

**CONCLUSIONS:** MetaVidence integrates the main forms of meta-analysis into a single validated interface that does not require programming. These findings support its use as an accessible and reproducible resource for evidence synthesis, particularly for researchers without programming skills or statistical support.

## BACKGROUND

Systematic reviews and meta-analyses are central to evidence-based medicine. Their validity, however, depends on the statistical methods used and on the transparent reporting of analytical decisions ^(1, 2)^. R provides a comprehensive environment for these analyses, with the meta ^(3)^ and metafor ^(4)^ packages supporting a wide diversity of effect measures, fixed-effect and random-effects models, prediction intervals and secondary analyses, and netmeta ^(5)^ extending these capabilities to network meta-analysis. Nevertheless, these packages require expertise in R syntax and package-specific data structures, which may limit their use by researchers without coding experience.

Most of the graphical tools available aim to address a single design, such as network meta-analysis ^(6)^, diagnostic test accuracy ^(7, 8)^, or combinations of pairwise, single-arm and, in some cases, network or diagnostic analyses ^(9-12)^. To our knowledge, no available tool performs pairwise, single-arm, pre-calculated, network and diagnostic test accuracy meta-analysis together. Thus, researchers whose reviews span several designs may have to move between programs and transcribe results manually, inviting errors. Moreover, these barriers weigh most on students, early-career investigators and research groups with limited funding or statistical support, including those in low- and middle-income countries ^(13)^.

In this study, we developed MetaVidence, a free and open-source no-code application that performs five families of meta-analysis within a single interface, with the aim of making established statistical methods accessible to a broader range of researchers. MetaVidence adapts well-established methods to a common graphical workflow, with secondary analyses, analysis of multiple outcomes and publication-ready outputs. Here, we describe the application and its validation against the underlying R packages, published meta-analyses and its intended users.

## METHOD DETAILS

### Implementation and workflow

We developed MetaVidence as a Shiny application freely available at https://metavidence.com and as the open-source R package metavidence. All modules follow the same three-step workflow of data input and verification, specification of the analytical settings and presentation of results (Figure 1).

**Figure 1.**
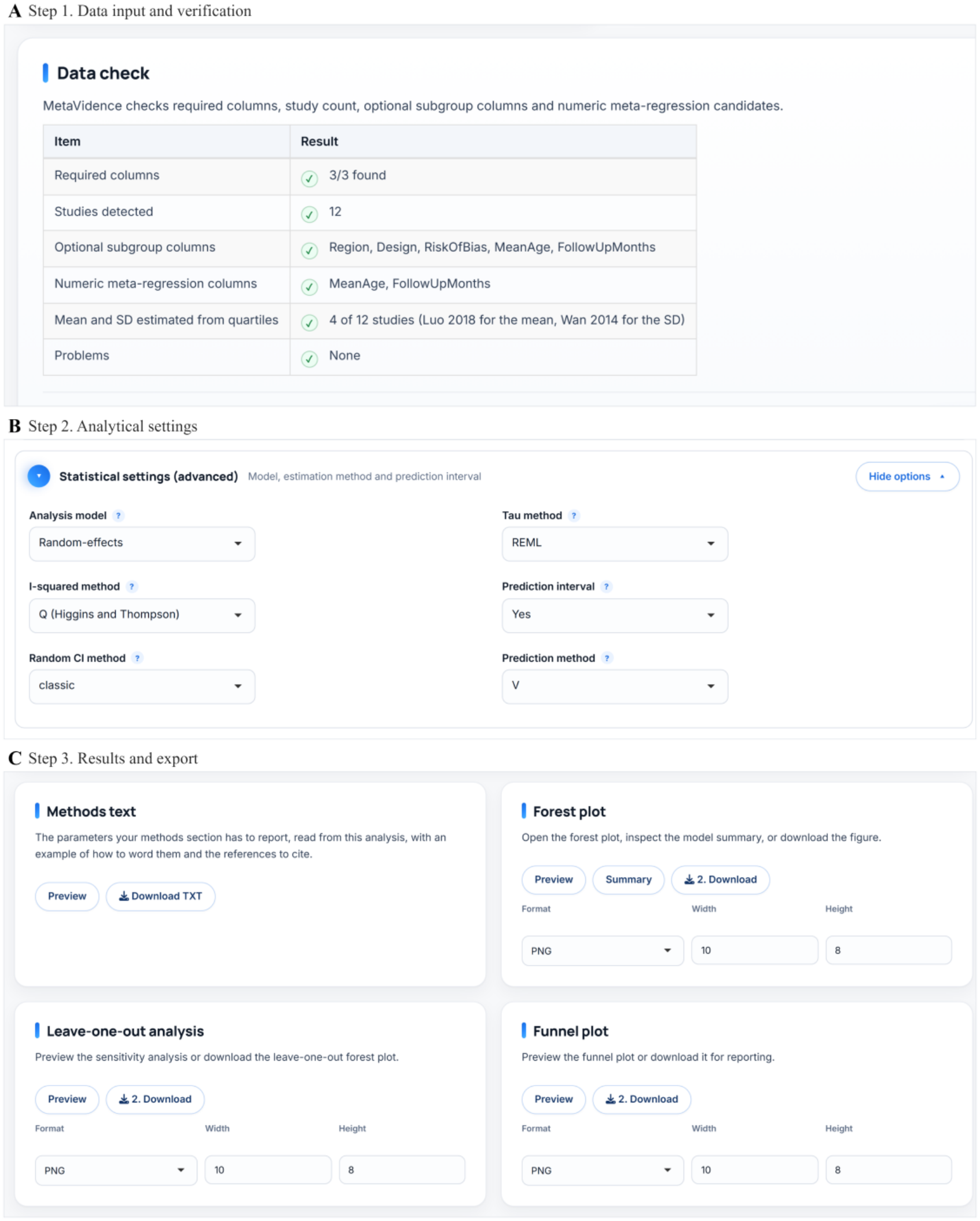
The three-step workflow shared by all MetaVidence modules, shown for the example continuous-outcome dataset. (A) Step 1, the data check after upload, reporting the required columns, the number of studies, the variables available for subgroup analysis and meta-regression, and the four studies whose means and standard deviations were estimated from medians and quartiles; (B) Step 2, the statistical settings, with the analysis model, between-study variance estimator, I^2^ method, confidence interval method and prediction interval; (C) Step 3, the result cards, each with its own preview, summary and download options.

#### Step 1. Data input and verification

Users can paste data directly from a spreadsheet or upload it as a .csv or .xlsx file, based on downloadable templates that define the variables required by each kind of analysis and allow the inclusion of study-level variables for subgroup analysis and meta-regression. Before the analytical settings are displayed, the application verifies the dataset and identifies missing columns, empty study labels, nonnumeric values, event counts exceeding sample sizes, invalid confidence limits, inconsistent treatment or study-arm coding and datasets with fewer than two studies. In addition, the data report identifies the variables available for secondary analyses and the studies in which means and standard deviations were estimated from medians and quartiles (Figure 1A).

#### Step 2. Analytical settings

Default values are provided for the most commonly used analyses. Alternative effect measures, models, heterogeneity estimators and inferential methods are also available (Figure 1B). Combinations not supported by the underlying packages are not offered to avoid errors.

#### Step 3. Results and export

Numerical estimates and graphical outputs are generated from the same fitted object. Each output is presented in a separate card that can be previewed, summarized or downloaded, with the file format and figure dimensions defined on the card itself (Figure 1C).

#### Deployment

The web and local versions are generated from the same source code. The web version is compiled with shinylive and runs through WebAssembly, and therefore all calculations are performed on the user’s device and study data are not transmitted to an external server. Similarly, the R package provides the same interface for offline use.

### Analysis families and statistical methods

MetaVidence comprises five analysis families. Pairwise analysis accepts event counts and sample sizes from two groups, means and standard deviations, or medians with quartiles or ranges, including datasets in which the reporting format differs across studies. Single-arm analysis includes proportions and means, and the pre-calculated family accepts study estimates reported with confidence intervals, standard errors, or both. The network family accepts arm-level binary and continuous data as well as contrast-level estimates, and diagnostic test accuracy can be analyzed for a single index test or for two tests evaluated in the same studies.

The pairwise, single-arm and pre-calculated families are built on meta ^(3)^ and metafor ^(4)^. Binary outcomes can be expressed as risk ratios, odds ratios or risk differences, and continuous outcomes as mean or standardized mean differences. Single proportions can be analyzed on raw or transformed scales and single means on the original or logarithmic scale, whereas hazard ratios and other relative or absolute measures are entered pre-calculated. Users can select fixed-effect or random-effects models, as well as the pooling method and the definition of I^2^. For random-effects analyses, the between-study variance estimators implemented in meta ^(3)^, alternative confidence intervals and prediction intervals are also available. When continuous outcomes are reported as medians, means are estimated with the method of Luo et al. ^(14)^, and standard deviations with the method of Wan et al. ^(15)^ when only the interquartile range or the range is reported, or with that of Shi et al. ^(16)^ when both are reported.

Network meta-analysis is performed with netmeta ^(5)^ under a frequentist graph-theoretical framework ^(17)^. Diagnostic test accuracy is analyzed with a bivariate binomial generalized linear mixed model ^(18)^ fitted through lme4 ^(19)^, according to the Cochrane Handbook for Diagnostic Test Accuracy Reviews ^(20)^. This model jointly estimates sensitivity and specificity and provides the summary operating point with its confidence and prediction regions. When two index tests are evaluated in the same studies, both summary operating points are estimated, and differences in sensitivity and specificity are assessed with likelihood-ratio tests.

Secondary analyses are integrated into the applicable modules. Leave-one-out analysis evaluates the influence of each study, subgroup analysis reports the estimates for each subgroup with the test for subgroup differences, and meta-regression uses numeric study-level moderators. Funnel plots may be accompanied by Egger’s test when appropriate for the selected analysis. All secondary analyses use the same effect measure, model and heterogeneity settings as the main analysis.

### Outputs

The pairwise and single-arm families share the same graphical outputs, including forest plots for the main analysis, subgroup analysis and leave-one-out analysis, bubble plots for meta-regression and funnel plots for small-study effects (Figure 2A-E). Plot dimensions and colors can be customized, and figures can be exported as 600-dpi .png or vector .pdf files. Moreover, subgroup and meta-regression results for multiple variables are exported as.xlsx tables containing the pooled estimates, confidence intervals, numbers of studies and participants, heterogeneity statistics, tests for subgroup differences, regression coefficients, P values and R^2^ (Figure 2F, G).

**Figure 2.**
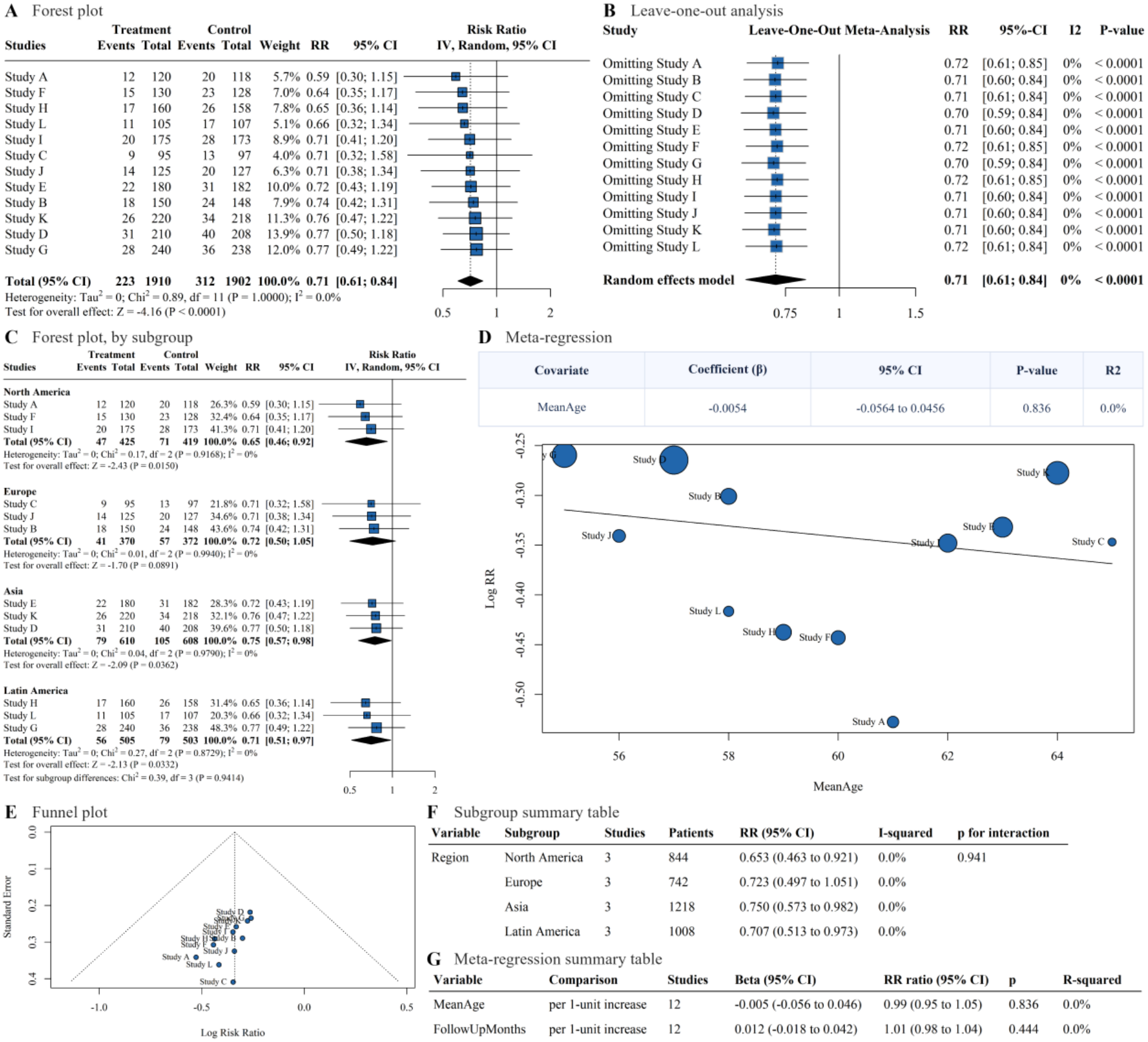
Pairwise and single-arm meta-analysis in MetaVidence, shown for a binary outcome. (A) Forest plot; (B) leave-one-out analysis; (C) the same forest plot with the subgroup structure and the test for subgroup differences; (D) meta-regression on a study-level moderator, with the coefficient table and the bubble plot; (E) funnel plot; (F) subgroup summary table; (G) meta-regression summary table.

The network family provides node-splitting results, network graphs, relative treatment effects, league tables and rankings by P-score (Figure 3A-C, E). Similarly, the diagnostic family provides study-level sensitivity and specificity with bivariate summary estimates in receiver operating characteristic space, including the comparison of two index tests evaluated in the same studies (Figure 3D, F).

**Figure 3.**
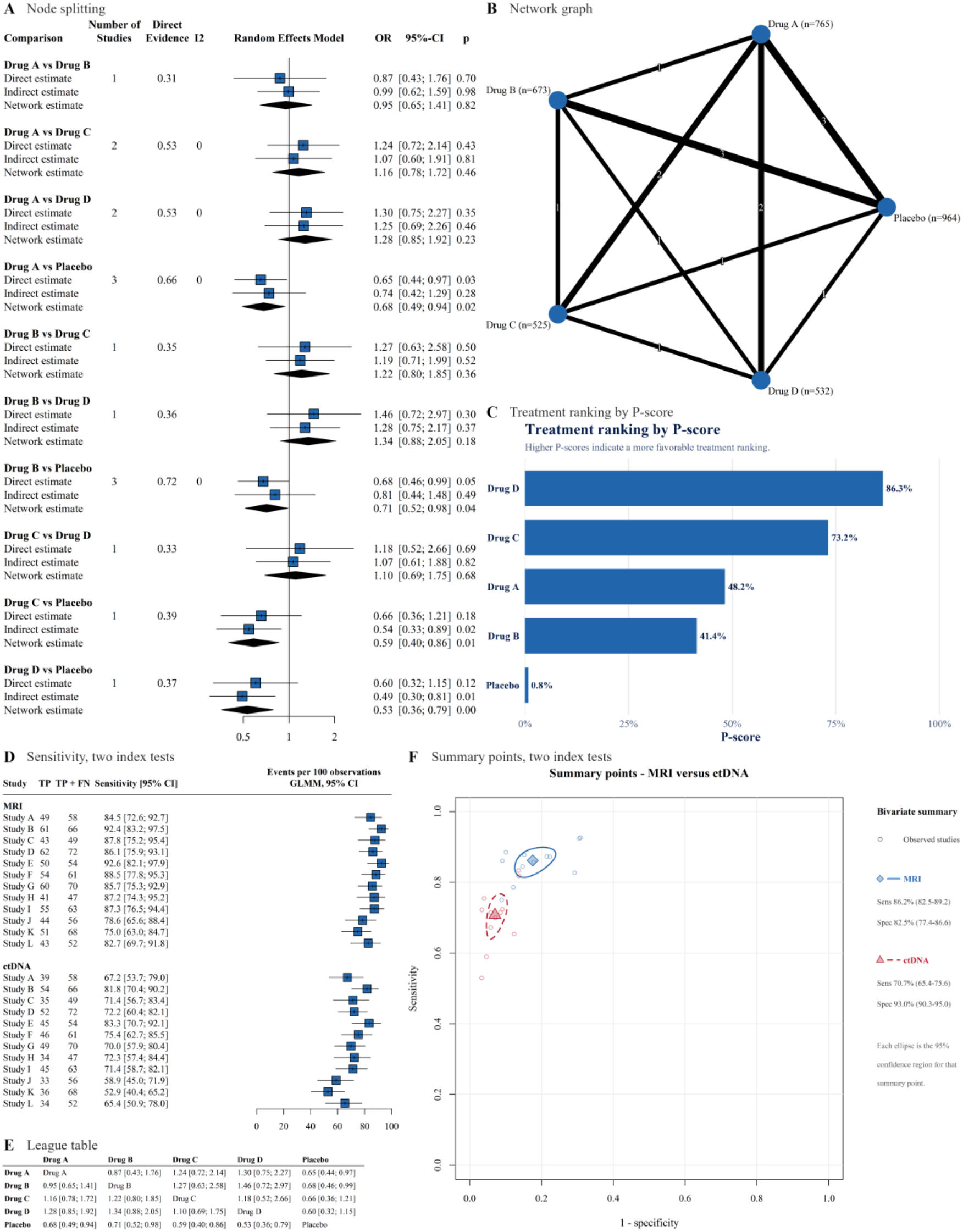
Network meta-analysis and diagnostic test accuracy in MetaVidence. (A) Node splitting of direct, indirect and network estimates; (B) network graph, with edge width scaled to the number of studies contributing to each comparison; (C) treatment ranking by P-score; (D) study-level sensitivity for two index tests evaluated in the same studies; (E) league table of all pairwise comparisons; (F) summary operating points for those two index tests from the bivariate model, each with its 95% confidence region.

### Analysis of multiple outcomes

Systematic reviews usually report several outcomes, which are often analyzed one at a time with the same specifications. Therefore, in the pairwise, single-arm and pre-calculated families, a workbook in which each sheet contains one outcome can be analyzed in a single run. The data check is applied to all sheets before any model is fitted, and the model, estimator, interval settings, subgroup variable and meta-regression moderator are selected once and applied to all outcomes. An overview reports, for each outcome, the number of studies and whether subgroup and meta-regression analyses were performed or skipped, with the corresponding reason. Each figure is exported as a compressed file with one image per outcome, and the subgroup and meta-regression tables of all outcomes are combined in a single spreadsheet.

### Reporting the analysis

The between-study variance estimator, the confidence interval method and the software version are frequently not reported in published meta-analyses ^(2)^. Thus, every analysis also generates a Methods text with two parts. The first part lists the parameters that should be reported in the methods section, extracted directly from the fitted model, including the summary measure, pooling method, model, between-study variance estimator, confidence and prediction interval methods, definition of I^2^, continuity correction, conversion of medians, secondary analyses and number of studies, as well as the versions of MetaVidence, R and the packages used. The second part provides an example of how these parameters may be reported, with the corresponding references.

## METHOD VALIDATION

### Computational agreement with the underlying packages

We evaluated whether the application correctly transferred the uploaded data and selected parameters to the underlying statistical packages. For this purpose, MetaVidence estimates were compared with results obtained from direct calls to meta ^(3)^, metafor ^(4)^ and netmeta ^(5)^, using the same datasets and statistical arguments. Comparisons included reference datasets, effect measures, model specifications, between-study variance estimators, confidence interval methods, definitions of I^2^, subgroup models, meta-regression, leave-one-out analyses, Egger’s test, network-specific outputs, batch processing and median-to-mean conversion. Validation was performed with meta 8.3.0, metafor 4.8.0 and netmeta 3.2.0.

A total of 6,321 checks were performed (Table S1). The maximum absolute numerical difference observed in the reference datasets was 3.9 × 10^−16^, which is consistent with floating-point precision.

The diagnostic module was validated separately, since the bivariate binomial model has no equivalent implementation in the packages used for the other designs. Thus, MetaVidence was applied to the anti-CCP example of 37 studies distributed with the Cochrane Handbook for Diagnostic Test Accuracy Reviews ^(20)^. The summary sensitivity (0.658) and specificity (0.957) agreed with the published estimates to within 0.0002.

### Reproduction of published meta-analyses

To evaluate the complete sequence from study-level data to reported results, we reconstructed 12 published meta-analyses, two from each of six input formats covering four of the five analysis families ^(21-32)^. These included pairwise binary and continuous outcomes, single proportions, single means, pre-calculated effects and diagnostic test accuracy. Meta-analyses were selected based on the availability of complete study-level data and adequately reported specifications, and agreement was judged against the precision reported in each publication. Network functions were evaluated separately with datasets distributed with netmeta ^(5)^.

All 12 meta-analyses were successfully reproduced, including binary outcomes (Figure S1), continuous outcomes (Figure S2), single proportions (Figure S3), single means (Figure S4), pre-calculated effects (Figure S5) and diagnostic test accuracy (Figures S6 and S7). All pooled estimates and confidence limits were reproduced within half of the last reported digit. The source publications, the settings used and both sets of estimates are presented in Table 1 ^(21-32)^.

**Table 1.** The twelve published meta-analyses reproduced in MetaVidence, with the settings used. The settings used for each reconstruction are listed, and agreement was assessed against the numerical precision reported in each source publication. Study-level input data are given in Supplementary Datasets S1-S12, one table per meta-analysis.

| Source | Outcome | Design | Settings used in MetaVidence | Published estimate (95% CI) | MetaVidence estimate (95% CI) | Published I², % | MetaVidence I², % |
|---|---|---|---|---|---|---|---|
| Costa Filho et al. <sup>(21)</sup> | Pathological complete response, % | Single proportion | Raw proportion, IV, random-effects, REML, classic CI | 47.78 (39.82 to 55.74) | 47.78 (39.82 to 55.74) | 13.3 | 13.3 |
| Dacoregio et al. <sup>(22)</sup> | Grade 3 or higher adverse events, % | Single proportion | Logit proportion, GLMM, random-effects, ML | 43.98 (28.61 to 60.60) | 43.98 (28.61 to 60.60) | 97 | 97 |
| Fonseca et al. <sup>(23)</sup> | Excess weight loss at 12 months | Single mean | Raw mean, IV, random-effects, REML, classic CI | 62.81 (52.56 to 73.07) | 62.81 (52.56 to 73.07) | 78 | 78 |
| Veeresh et al. <sup>(24)</sup> | Cerebellar tonsil position | Single mean | Raw mean, IV, random-effects, DL, Hartung-Knapp CI | 1.13 (0.51 to 1.76) | 1.13 (0.51 to 1.76) | 98.1 | 98.1 |
| Defante et al. <sup>(25)</sup> | Pain at 15 days postpartum | Binary outcome | RR, Mantel-Haenszel, random-effects, DL, classic CI | RR 0.74 (0.65 to 0.83) | RR 0.74 (0.65 to 0.83) | 56.9 | 56.9 |
| Katsuyama et al. <sup>(26)</sup> | Any reoperation | Binary outcome | OR, Mantel-Haenszel, random-effects, REML, classic CI | OR 0.73 (0.60 to 0.89) | OR 0.73 (0.60 to 0.89) | 0 | 0 |
| Mamede et al. <sup>(27)</sup> | Vasopressin duration | Continuous outcome | MD, IV, random-effects, REML, classic CI | MD -8.77 (-21.67 to 4.13) | MD -8.77 (-21.67 to 4.13) | 17.9 | 17.9 |
| Ferreira et al. <sup>(28)</sup> | Days of mechanical ventilation | Continuous outcome | MD, IV, random-effects, DL, classic CI | MD -4.34 (-6.98 to -1.69) | MD -4.34 (-6.98 to -1.69) | 87 | 87.2 |
| Noronha et al. <sup>(29)</sup> | Overall survival | Pre-calculated effect | HR, IV, random-effects, REML, classic CI | HR 3.32 (2.09 to 5.29) | HR 3.32 (2.09 to 5.29) | 48 | 47.5 |
| Lee et al. <sup>(30)</sup> | Overall survival | Pre-calculated effect | HR, IV, random-effects, DL, classic CI | HR 1.48 (1.24 to 1.76) | HR 1.48 (1.24 to 1.76) | 34.1 | 34 |
| Fomenko et al. <sup>(31)</sup> | Sensitivity of HADS-A | Diagnostic accuracy | Bivariate binomial GLMM | 0.74 (0.70 to 0.78) | 0.739 (0.697 to 0.776) |  |  |
| Fomenko et al. <sup>(31)</sup> | Specificity of HADS-A | Diagnostic accuracy | Bivariate binomial GLMM | 0.76 (0.73 to 0.79) | 0.764 (0.734 to 0.792) |  |  |
| Akturk et al. <sup>(32)</sup> | Sensitivity of GAD-2 | Diagnostic accuracy | Bivariate binomial GLMM | 0.68 (0.59 to 0.75) | 0.675 (0.594 to 0.748) |  |  |
| Akturk et al. <sup>(32)</sup> | Specificity of GAD-2 | Diagnostic accuracy | Bivariate binomial GLMM | 0.86 (0.82 to 0.89) | 0.857 (0.820 to 0.888) |  |  |

### Usability

Usability was assessed in a cross-sectional, unsupervised study. Participants were recruited through social media and through email invitations sent to corresponding authors of recently published meta-analyses. Researchers who had previously conducted or published at least one meta-analysis were eligible, with no other restriction, and no incentive was offered.

Participants received written instructions to perform a meta-analysis of a binary outcome using a provided dataset, and were then invited to freely explore the remaining modules before completing an anonymous online questionnaire (Appendix S1). The questionnaire included the ten items of the System Usability Scale ^(33)^, scored on a five-point agreement scale, followed by items on the number of meta-analyses previously conducted, time and ease of use compared with R and RevMan, the impact of analyzing several outcomes in one run, intention of future use and likelihood of recommendation. Scores were calculated according to the standard method, in which one point is subtracted from each positively worded item, each negatively worded item is subtracted from five, and the sum is multiplied by 2.5, resulting in a score from 0 to 100. Responses with any missing scale item were excluded. The mean score is reported with a 95% confidence interval based on the t distribution, and the remaining items are reported as proportions.

A total of 60 responses were received, of which 59 contained all ten scale items. All respondents had previously conducted or published at least one meta-analysis (median 4, IQR 3-6). The mean System Usability Scale score was 82.8 (95% CI 76.2-89.5), above the benchmark mean of 68 for software in general and for digital health applications ^(34)^. Responses to each item are shown in Figure S8.

Compared with their previous use of R, 88% of participants considered the analytical process shorter or much shorter, and 88% considered MetaVidence easier or much easier to use. Similarly, compared with RevMan, 83% reported a shorter process and 80% reported greater ease of use (Figure S9). Regarding the analysis of several outcomes at once, 80% reported that it made their workflow considerably or much faster, and 93% reported some degree of gain. Moreover, 78% would probably or definitely use MetaVidence in a future review, and 76% rated their likelihood of recommending it as 9 or 10 on a 10-point scale.

## LIMITATIONS

The strengths of our study include the validation of MetaVidence against direct package calls, published meta-analyses and a reference diagnostic example, as well as its evaluation by researchers with previous experience in meta-analysis. Nonetheless, our study has limitations. The usability evaluation included a self-selected sample and was conducted without supervision, so its findings may not apply equally to inexperienced users. Moreover, perceived ease and speed do not establish improvements in analytical accuracy or efficiency, and the time saved with the analysis of multiple outcomes was reported by participants instead of measured. Finally, MetaVidence supports only the statistical stage of evidence synthesis, and methodological training remains essential for the choice of appropriate analyses and their correct interpretation.

## CONCLUSION

Our study showed that MetaVidence agrees with the underlying R packages and reproduces published meta-analyses, while integrating pairwise, single-arm, pre-calculated, network and diagnostic test accuracy meta-analysis into a free interface that does not require programming. These findings support MetaVidence as an accessible and reproducible resource for evidence synthesis, which may enable a broader range of researchers to conduct the statistical stage of systematic reviews.

## ETHICS STATEMENT

The usability study involved adult researchers responding to an anonymous online questionnaire. No name, email address, institution or other identifying information was collected, and responses were analyzed only in aggregate. Participation was voluntary and unpaid, with no coercion to participate, and participants were informed before responding of the purpose of the study and that the aggregated results could be published in a scientific article; completing the questionnaire after reading this information constituted consent to participate. Under Brazilian National Health Council Resolution CNS 510/2016 (Article 1, sole paragraph, item I), anonymous opinion research is exempt from review by a research ethics committee.

## AUTHOR CONTRIBUTIONS

**Valbert Oliveira Costa Filho:** Conceptualization, Methodology, Software, Validation, Formal analysis, Investigation, Data curation, Writing - original draft, Writing - review and editing, Visualization. **Pedro Robson Costa Passos:** Data curation, Validation, Writing - review and editing. **Mariana Macambira Noronha:** Data curation, Validation, Writing - review and editing. **Rafael Lara Nohmi:** Validation, Writing - review and editing. **Luis Felipe Leite da Silva:** Validation, Writing - review and editing. **Erick F. Saldanha:** Writing - review and editing. **Giuseppe G. F. Leite:** Software, Validation, Writing - review and editing.

## DATA AVAILABILITY

MetaVidence is freely available at https://metavidence.com. Step-by-step tutorials are available at https://metavidence.com/tutorials/en/, and the source code is open source under the GPL-3.0 license. Study-level input data of the reproduced meta-analyses (Datasets S1-S12) are provided as supplementary material.

## SUPPLEMENTARY MATERIAL

Appendix S1 (usability questionnaire); Table S1 (computational validation checks); Figures S1-S7 (reproduced meta-analyses); Figures S8 and S9 (usability responses); Datasets S1-S12 (study-level input data).

## REFERENCES

1. Kanukula R, Page MJ, Turner SL, McKenzie JE. Identification of application and interpretation errors that can occur in pairwise meta-analyses in systematic reviews of interventions: a systematic review. Journal of clinical epidemiology. 2024;170:111331.

2. Arredondo Montero J. Reporting and interpreting meta-analysis software choices: practical guidance for researchers, reviewers, and editors. European journal of clinical pharmacology. 2026;82(8):205.

3. Balduzzi S, Rucker G, Schwarzer G. How to perform a meta-analysis with R: a practical tutorial. Evidence-based mental health. 2019;22(4):153–60.

4. Viechtbauer W. Conducting Meta-Analyses in R with the metafor Package. Journal of Statistical Software. 2010;36(3).

5. Balduzzi S, Rücker G, Nikolakopoulou A, Papakonstantinou T, Salanti G, Efthimiou O, et al. netmeta: An R Package for Network Meta-Analysis Using Frequentist Methods. Journal of Statistical Software. 2023;106(2).

6. Owen RK, Bradbury N, Xin Y, Cooper N, Sutton A. MetaInsight: An interactive web-based tool for analyzing, interrogating, and visualizing network meta-analyses using R-shiny and netmeta. Research synthesis methods. 2019;10(4):569–81.

7. Freeman SC, Kerby CR, Patel A, Cooper NJ, Quinn T, Sutton AJ. Development of an interactive web-based tool to conduct and interrogate meta-analysis of diagnostic test accuracy studies: MetaDTA. BMC medical research methodology. 2019;19(1):81.

8. Mizutani S, Zhou Y, Tian Y-S, Takagi T, Ohkubo T, Hattori S. DTAmetasa: An R shiny application for meta-analysis of diagnostic test accuracy and sensitivity analysis of publication bias. Research synthesis methods. 2023;14(6):916–25.

9. Wallace BC, Schmid CH, Lau J, Trikalinos TA. Meta-Analyst: software for meta-analysis of binary, continuous and diagnostic data. BMC medical research methodology. 2009;9:80.

10. Suurmond R, van Rhee H, Hak T. Introduction, comparison, and validation of Meta-Essentials: A free and simple tool for meta-analysis. Research synthesis methods. 2017;8(4):537–53.

11. Elmakaty I. Mastering meta-analysis in Microsoft Excel with MetaXL add-in: A comprehensive tutorial and guide to meta-analysis. Journal of evaluation in clinical practice. 2025;31(2):e14138.

12. Fekete JT, Gyorffy B. MetaAnalysisOnline.com: Web-Based Tool for the Rapid Meta-Analysis of Clinical and Epidemiological Studies. Journal of medical Internet research. 2025;27:e64016.

13. Oliver S, Bangpan M, Stansfield C, Stewart R. Capacity for conducting systematic reviews in low-and middle-income countries: a rapid appraisal. Health research policy and systems. 2015;13:23.

14. Luo D, Wan X, Liu J, Tong T. Optimally estimating the sample mean from the sample size, median, mid-range, and/or mid-quartile range. Statistical methods in medical research. 2018;27(6):1785–805.

15. Wan X, Wang W, Liu J, Tong T. Estimating the sample mean and standard deviation from the sample size, median, range and/or interquartile range. BMC Medical Research Methodology. 2014;14:135.

16. Shi J, Luo D, Weng H, Zeng X-T, Lin L, Chu H, et al. Optimally estimating the sample standard deviation from the five-number summary. Research synthesis methods. 2020;11(5):641–54.

17. Rucker G. Network meta-analysis, electrical networks and graph theory. Research synthesis methods. 2012;3(4):312–24.

18. Chu H, Cole SR. Bivariate meta-analysis of sensitivity and specificity with sparse data: a generalized linear mixed model approach. Journal of clinical epidemiology. 2006;59(12):1331–3.

19. Bates D, Mächler M, Bolker B, Walker S. Fitting Linear Mixed-Effects Models Using lme4. Journal of Statistical Software. 2015;67(1).

20. Takwoingi Y, Dendukuri N, Schiller I, Rücker G, Jones HE, Partlett C, et al. Undertaking meta-analysis. Cochrane Handbook for Systematic Reviews of Diagnostic Test Accuracy: Wiley; 2023. p. 249–325.

21. Filho VOC, Noronha MM, Passos PRC, Ribeiro MF, Saldanha EF. Neoadjuvant Immune Checkpoint Inhibitors in CSCC: A Systematic Review and Meta-Analysis. Journal of cutaneous medicine and surgery. 2026;30(2):175–6.

22. Dacoregio MI, Rego Castro CEd, Mamede I, Rodrigues CG, Thimotheo Batista JP, Michelon I, et al. Toxicity profile and tolerability of immune checkpoint inhibitors for the treatment of early-stage breast cancer: a systematic review and meta-analysis. Breast (Edinburgh, Scotland). 2026;88:104821.

23. Fonseca P, Pereira L, Braga JG, Scremin GM, de Araujo L, Alves J, et al. Efficacy and Safety of Bariatric Surgery in Well-Compensated Liver Cirrhosis: A Systematic Review and a Single-Arm Meta-analysis. Obesity surgery. 2025;35(10):4246–63.

24. Veeresh V, Hosapatna M, Menon G, Shekar M, Pruthi N, Kausar A. Normative cerebellar tonsil position in healthy individuals: a systematic review and meta-analysis. European spine journal : official publication of the European Spine Society, the European Spinal Deformity Society, and the European Section of the Cervical Spine Research Society. 2026;35(6):2977–84.

25. Defante MLR, de Souza K, Oliveira JA, Inoue Salgado AS, de Oliveira FR, Cardoso FDS. Continuous versus interrupted suturing for second-degree perineal tears and episiotomies: A meta-analysis of randomized trials. European journal of obstetrics, gynecology, and reproductive biology. 2025;314:114722.

26. Katsuyama E, Fukunaga C, Passos FS, Lee N, Ventura de Santana de Jesus AC, Ydy CM, et al. Long-term outcomes of surgical repair versus replacement for tricuspid valve endocarditis -A meta-analysis of reconstructed time-to-event data. International journal of cardiology Heart & vasculature. 2025;60:101782.

27. Mamede I, Area L, Carvalhal G, Bessa R, Lenzi M, Dos Santos MCF. Early vasopressin plus norepinephrine versus delayed or no vasopressin in septic shock: A systematic review and meta-analysis. The American journal of emergency medicine. 2026;99:225–31.

28. Ferreira ROM, Pasqualotto E, Viana P, Schmidt PHS, Andrighetti L, Chavez MP, et al. Surgical versus non-surgical treatment of flail chest: a meta-analysis of randomized controlled trials. European journal of trauma and emergency surgery : official publication of the European Trauma Society. 2023;49(6):2531–41.

29. Noronha MM, da Silva LFL, Almeida LFC, Passos PRC, Reis PCA, Conrado JEP, et al. Prognostic and Predictive Value of ctDNA for Metastatic Uveal Melanoma: A Systematic Review and Meta-Analysis. Pigment cell & melanoma research. 2025;38(5):e70047.

30. Lee S, Correa TL, Birda V, Roshan K, Peters ML, Modest AM, et al. Impact of prior conventional irinotecan use on nanoliposomal irinotecan efficacy in advanced pancreatic cancer: a meta-analysis. ESMO open. 2026;11(8):107774.

31. Fomenko A, Dummler D, Akturk Z, Eck S, Teusen C, Karapetyan S, et al. Hospital Anxiety and Depression Scale Anxiety subscale (HADS-A) for detecting anxiety disorders in adults. The Cochrane database of systematic reviews. 2025;7(7):CD015456.

32. Akturk Z, Hapfelmeier A, Fomenko A, Dummler D, Eck S, Olm M, et al. Generalized Anxiety Disorder 7-item (GAD-7) and 2-item (GAD-2) scales for detecting anxiety disorders in adults. The Cochrane database of systematic reviews. 2025;3(3):CD015455.

33. Brooke J. SUS: a quick and dirty usability scale. In: Jordan PW, Thomas B, Weerdmeester BA, McClelland IL, editors. Usability Evaluation in Industry. London: Taylor and Francis; 1996. p. 189–94.

34. Hyzy M, Bond R, Mulvenna M, Bai L, Dix A, Leigh S, et al. System Usability Scale Benchmarking for Digital Health Apps: Meta-analysis. JMIR mHealth and uHealth. 2022;10(8):e37290.

