## Supplementary Material for "Development and Validation of MetaVidence: A No-Code Application for Accessible and Reproducible Meta-analysis"

| Check | What is compared | Comparisons | Result |
|---|---|---|---|
| Reference datasets | One published reference dataset per module, run through the application, against a direct call to meta, metafor or netmeta | 36 | Maximum absolute difference 3.9e-16 |
| Option sweep | Full cross of effect measure, model, between-study variance estimator, confidence interval method and I-squared definition, against the same call to meta | 5,622 | No divergence |
| Option sweep (concordant refusals) | Combinations the application and the package both refuse, for the same reason | 255 | All concordant |
| Secondary analyses | Subgroup estimates, meta-regression coefficients, leave-one-out estimates and Egger p values | 252 | All identical |
| Network-specific analyses | Node splitting, P-scores and the league table, against netmeta | 42 | All identical |
| Batch equivalence | Each sheet of a multi-outcome workbook against the same sheet analyzed on its own | 96 | All identical |
| Median-to-mean conversion | Converted mean and standard deviation against the published formulae, and against the values reaching the pooled estimate | 18 | All identical |
| Total | | 6,321 | |

**Table S1.** The automated validation program, by class of check.

**A**

| Studies | Experimental Events | Experimental Total | Control Events | Control Total | Weight | RR | 95% CI |
|---|---|---|---|---|---|---|---|
| Iqbal | 28 | 100 | 57 | 100 | 7.2% | 0.49 | [0.34; 0.70] |
| Morano | 32 | 107 | 58 | 107 | 7.6% | 0.55 | [0.39; 0.77] |
| Martinez-Galiano | 22 | 70 | 34 | 64 | 5.9% | 0.59 | [0.39; 0.90] |
| Kettle | 204 | 771 | 338 | 771 | 14.5% | 0.60 | [0.52; 0.70] |
| Isager-Sally | 156 | 262 | 214 | 261 | 15.5% | 0.73 | [0.65; 0.81] |
| Mahomed | 129 | 553 | 150 | 524 | 12.1% | 0.81 | [0.67; 1.00] |
| Almeida | 15 | 48 | 18 | 47 | 3.9% | 0.82 | [0.47; 1.42] |
| Croce | 40 | 100 | 50 | 102 | 8.4% | 0.82 | [0.60; 1.11] |
| Kindberg | 65 | 200 | 72 | 200 | 9.6% | 0.90 | [0.69; 1.19] |
| Sadat-Un-Nisa | 21 | 42 | 38 | 76 | 6.7% | 1.00 | [0.69; 1.46] |
| Valenzuela | 42 | 223 | 41 | 222 | 6.5% | 1.02 | [0.69; 1.50] |
| Perveen | 11 | 100 | 10 | 100 | 2.1% | 1.10 | [0.49; 2.47] |
| **Total (95% CI)** | **765** | **2576** | **1080** | **2574** | **100.0%** | **0.74** | **[0.65; 0.83]** |

Heterogeneity: $Tau^2 = 0.0221$; $Chi^2 = 25.49$, df = 11 (P = 0.0077); $I^2 = 56.9\%$

Test for overall effect: Z = -4.87 (P < 0.0001)

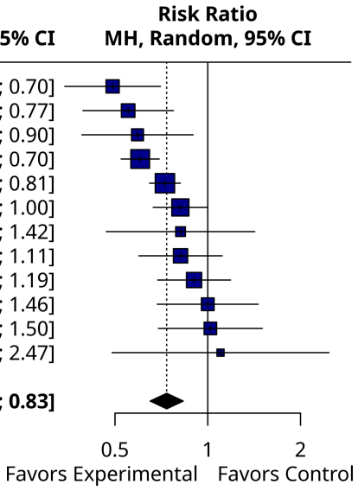


**B**

| Studies | Experimental Events | Experimental Total | Control Events | Control Total | Weight | OR | 95% CI |
|---|---|---|---|---|---|---|---|
| Xie, 2023 | 0 | 23 | 6 | 33 | 0.4% | 0.09 | [0.00; 1.68] |
| Shetty, 2015 | 0 | 5 | 1 | 2 | 0.3% | 0.09 | [0.00; 3.59] |
| Brescia, 2022 | 0 | 37 | 3 | 34 | 0.4% | 0.12 | [0.01; 2.41] |
| Pfannmueller, 2015 | 1 | 34 | 3 | 22 | 0.7% | 0.19 | [0.02; 1.98] |
| Baraki, 2013 | 1 | 15 | 2 | 18 | 0.6% | 0.57 | [0.05; 7.00] |
| Slaughter, 2019 | 48 | 532 | 130 | 962 | 26.8% | 0.63 | [0.45; 0.90] |
| Gaga, 2013 | 39 | 354 | 75 | 490 | 19.7% | 0.69 | [0.45; 1.04] |
| Lee, 2020 | 23 | 412 | 21 | 292 | 9.5% | 0.76 | [0.41; 1.41] |
| Thourani, 2024 | 70 | 1177 | 248 | 3654 | 40.8% | 0.87 | [0.66; 1.14] |
| Protos, 2018 | 1 | 12 | 1 | 26 | 0.4% | 2.27 | [0.13; 39.73] |
| Renzulli, 1999 | 1 | 11 | 0 | 10 | 0.3% | 3.00 | [0.11; 82.40] |
| **Total (95% CI)** | **184** | **2612** | **490** | **5543** | **100.0%** | **0.73** | **[0.60; 0.89]** |

Heterogeneity: $Tau^2 = 0.0039$; $Chi^2 = 9.45$, df = 10 (P = 0.4901); $I^2 = 0.0\%$

Test for overall effect: Z = -3.19 (P = 0.0014)

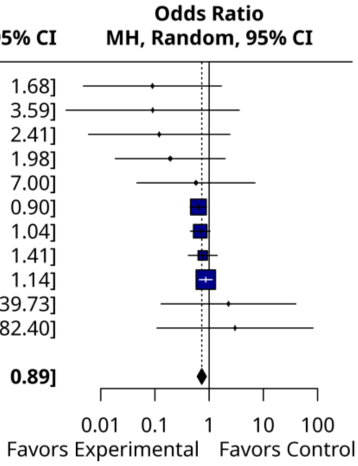


**Figure S1.** Binary outcomes reproduced in MetaVidence. (A) Defante et al.; (B) Katsuyama et al.

**A**

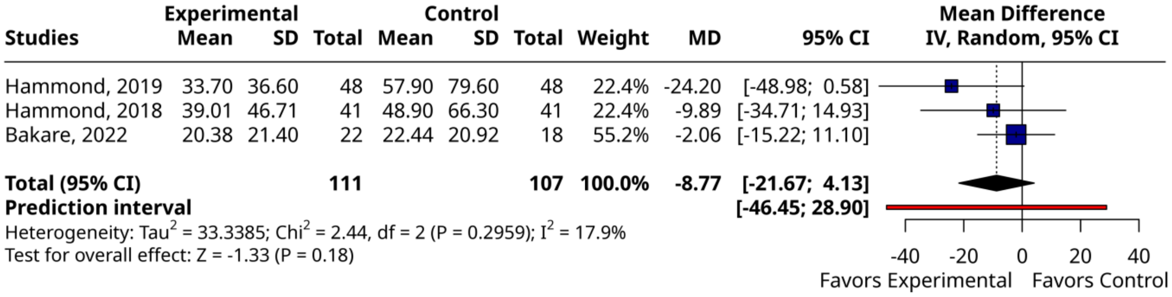


**B**

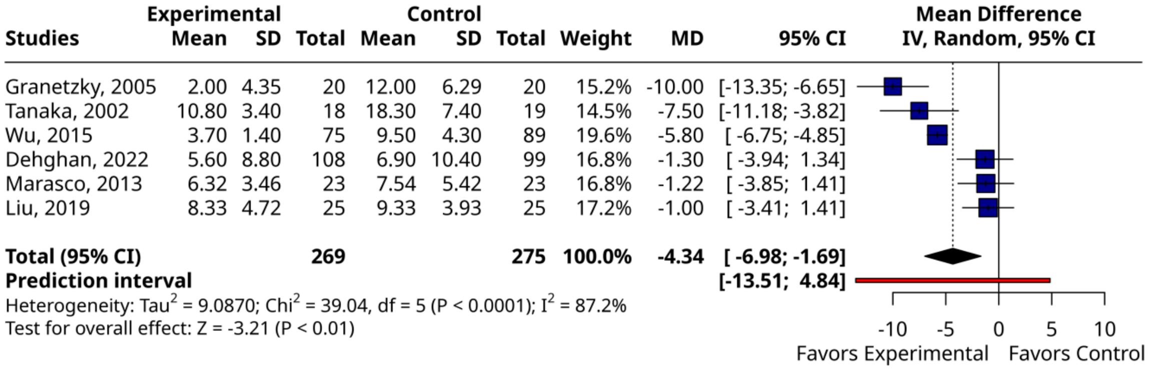


**Figure S2.** Continuous outcomes reproduced in MetaVidence. (A) Mamede et al.; (B) Ferreira et al.

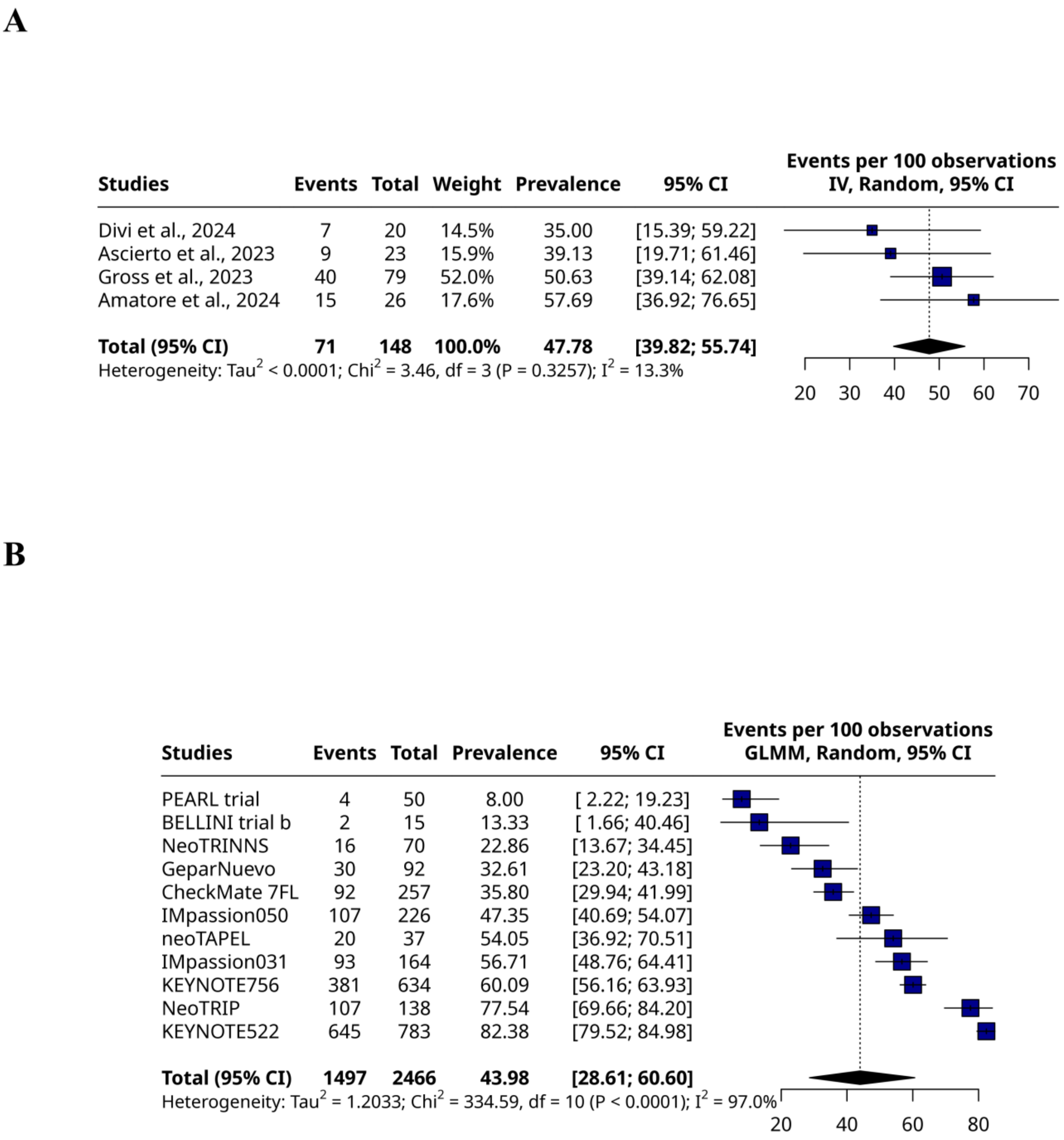


**Figure S3.** Single proportions reproduced in MetaVidence. (A) Costa Filho et al.; (B) Dacoregio et al.

**A**

| Studies | Mean | SD | Total | Weight | Mean | 95% CI |
|---|---|---|---|---|---|---|
| Hanipah et al., 2018 | 42.24 | 21.40 | 11 | 14.7% | 42.24 | [29.59; 54.89] |
| Sharpton et al., 2019 | 50.75 | 20.80 | 24 | 16.8% | 50.75 | [42.43; 59.07] |
| Woodford et al., 2015 | 61.30 | 24.10 | 11 | 13.8% | 61.30 | [47.06; 75.54] |
| Dalal et al., 2004 | 62.70 | 14.90 | 30 | 18.0% | 62.70 | [57.37; 68.03] |
| Rebibo et al., 2014 | 74.43 | 23.56 | 5 | 10.7% | 74.43 | [53.78; 95.08] |
| Garcia-Sesma et al., 2019 | 76.30 | 27.55 | 8 | 11.4% | 76.30 | [57.21; 95.39] |
| Mehdorn et al., 2022 | 79.81 | 33.55 | 27 | 14.7% | 79.81 | [67.16; 92.46] |
| **Total (95% CI)** | | | **116** | **100.0%** | **62.81** | **[52.56; 73.07]** |
| **Prediction interval** | | | | | | **[30.67; 94.96]** |

Heterogeneity: $Tau^2 = 145.1603$; $Chi^2 = 27.24$, df = 6 (P = 0.0001); $I^2 = 78.0\%$

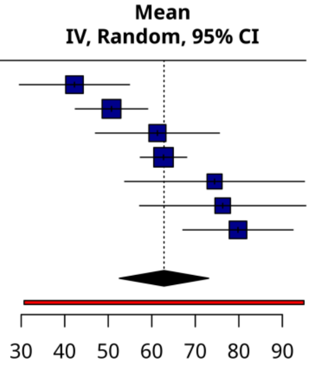


**B**

| Studies | Mean | SD | Total | Weight | Mean | 95% CI |
|---|---|---|---|---|---|---|
| Brandon W. Smith 2013 | -1.01 | 3.22 | 2400 | 7.2% | -1.01 | [-1.14; -0.88] |
| David J. Mikulis 1992 | -0.06 | 1.78 | 221 | 7.2% | -0.06 | [-0.29; 0.17] |
| Daniel P.M. Barros 2022 | 0.23 | 2.33 | 30 | 6.6% | 0.23 | [-0.60; 1.06] |
| Garland EM 2002 | 0.40 | 2.60 | 46 | 6.7% | 0.40 | [-0.35; 1.15] |
| Braden J. Lawrence 2018 | 0.48 | 2.88 | 10 | 4.9% | 0.48 | [-1.31; 2.27] |
| A.H. Aiken 2012 | 0.70 | 1.90 | 44 | 6.9% | 0.70 | [ 0.14; 1.26] |
| Ali Riza Guvercin 2023 | 0.89 | 1.95 | 250 | 7.2% | 0.89 | [ 0.65; 1.13] |
| Sheng Yang 2021 | 0.99 | 2.32 | 180 | 7.1% | 0.99 | [ 0.65; 1.33] |
| A. J. Barkovich 1986 | 1.00 | 1.90 | 200 | 7.2% | 1.00 | [ 0.74; 1.26] |
| Andreea G. Moore 2016 | 1.50 | 3.00 | 110 | 6.9% | 1.50 | [ 0.94; 2.06] |
| Shamar J. Young 2018 | 1.98 | 2.45 | 200 | 7.1% | 1.98 | [ 1.64; 2.32] |
| Kazuhide Furuya 1998 | 2.10 | 2.80 | 9 | 4.8% | 2.10 | [ 0.27; 3.93] |
| Ryan K. L. Lee 2015 | 2.20 | 1.80 | 18 | 6.6% | 2.20 | [ 1.37; 3.03] |
| Aboulezz Ahmed O 1985 | 2.90 | 3.40 | 82 | 6.7% | 2.90 | [ 2.16; 3.64] |
| DIPANKAR BISWAS 2019 | 3.01 | 3.42 | 140 | 6.9% | 3.01 | [ 2.44; 3.58] |
| **Total (95% CI)** | | | **3940** | **100.0%** | **1.13** | **[ 0.51; 1.76]** |
| **Prediction interval** | | | | | | **[-1.74; 4.01]** |

Heterogeneity: $Tau^2 = 1.6740$; $Chi^2 = 731.52$, df = 14 (P < 0.0001); $I^2 = 98.1\%$

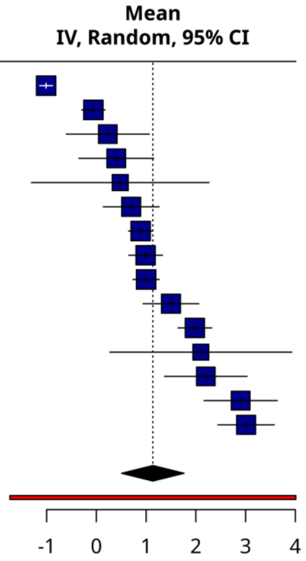


**Figure S4.** Single means reproduced in MetaVidence. (A) Fonseca et al.; (B) Veeresh et al.

**A**

| Studies | lnHR | SE | Weight | HR | 95% CI |
|---|---|---|---|---|---|
| Mariani, 2023 | 0.6366 | 0.4521 | 15.5% | 1.89 | [0.78; 4.59] |
| Hassel, 2023 | 0.7129 | 0.1742 | 30.0% | 2.04 | [1.45; 2.87] |
| Beasley, 2023 (cohort B) | 1.1378 | 0.7770 | 7.4% | 3.12 | [0.68; 14.30] |
| Carvajal, 2022 | 1.4702 | 0.2928 | 23.1% | 4.35 | [2.45; 7.72] |
| Ramelyte, 2024 | 1.5151 | 0.7623 | 7.6% | 4.55 | [1.02; 20.25] |
| Ny, 2021 | 1.8976 | 0.6827 | 9.0% | 6.67 | [1.75; 25.43] |
| Rodrigues, 2024 | 2.4078 | 0.7772 | 7.4% | 11.11 | [2.42; 50.92] |
| **Total (95% CI)** | | | **100.0%** | **3.32** | **[2.09; 5.29]** |

Heterogeneity: $Tau^2 = 0.1571$; $Chi^2 = 11.44$, df = 6 (P = 0.0758); $I^2 = 47.5\%$
Test for overall effect: Z = 5.07 (P < 0.0001)

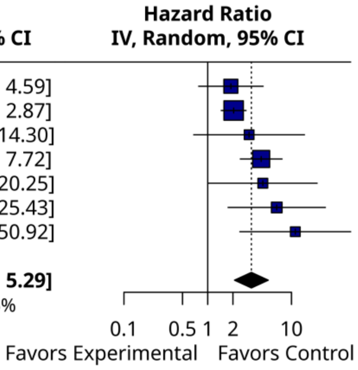


**B**

| Studies | lnHR | SE | Weight | HR | 95% CI |
|---|---|---|---|---|---|
| Kodama 2024 | -0.1278 | 0.3289 | 5.8% | 0.88 | [0.46; 1.67] |
| Harada 2023 | -0.0408 | 0.4106 | 4.0% | 0.96 | [0.43; 2.15] |
| Keum 2024 | 0.2390 | 0.1683 | 14.3% | 1.27 | [0.91; 1.76] |
| Yu 2023 | 0.2546 | 0.0962 | 21.8% | 1.29 | [1.07; 1.56] |
| Kasi 2021 | 0.2700 | 0.3018 | 6.7% | 1.31 | [0.72; 2.35] |
| Chun 2022 | 0.3436 | 0.2217 | 10.3% | 1.41 | [0.91; 2.17] |
| Chiu 2022 | 0.4762 | 0.1365 | 17.3% | 1.61 | [1.23; 2.10] |
| Kawakami 2023 | 0.4824 | 0.3993 | 4.2% | 1.62 | [0.74; 3.54] |
| Mohring 2023 | 0.6206 | 0.5445 | 2.4% | 1.86 | [0.64; 5.41] |
| Glassman 2018 | 0.8459 | 0.3213 | 6.0% | 2.33 | [1.24; 4.37] |
| Okada 2025 | 1.0006 | 0.4575 | 3.3% | 2.72 | [1.11; 6.67] |
| Yu 2022 | 1.4327 | 0.4228 | 3.8% | 4.19 | [1.83; 9.60] |
| **Total (95% CI)** | | | **100.0%** | **1.48** | **[1.24; 1.76]** |

Heterogeneity: $Tau^2 = 0.0267$; $Chi^2 = 16.68$, df = 11 (P = 0.1178); $I^2 = 34.0\%$
Test for overall effect: Z = 4.41 (P < 0.0001)

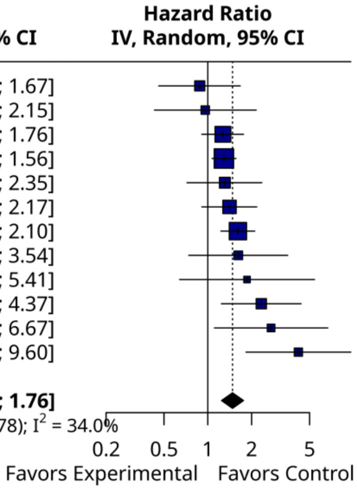


**Figure S5.** Pre-calculated effect sizes reproduced in MetaVidence. (A) Noronha et al.; (B) Lee et al.

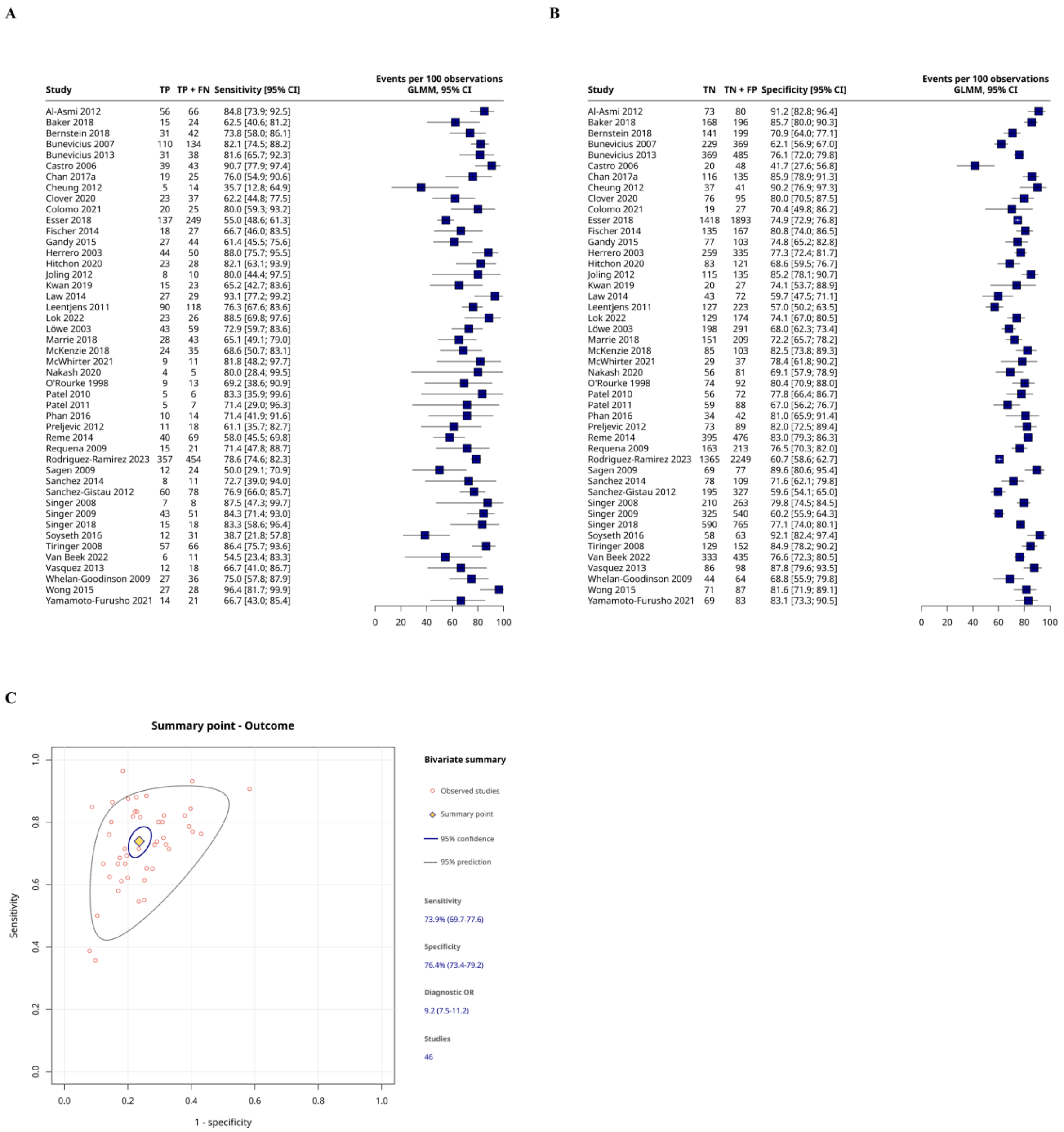


**Figure S6.** Diagnostic test accuracy reproduced in MetaVidence, Fomenko et al. (A) Sensitivity; (B) specificity; (C) bivariate summary point.

A

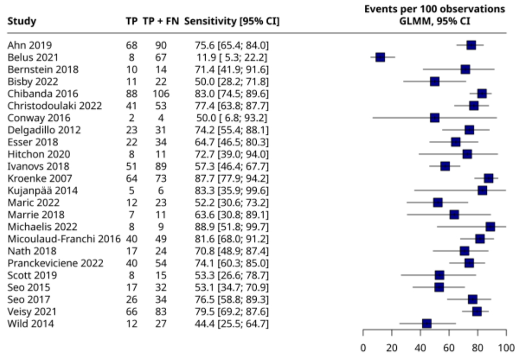


B

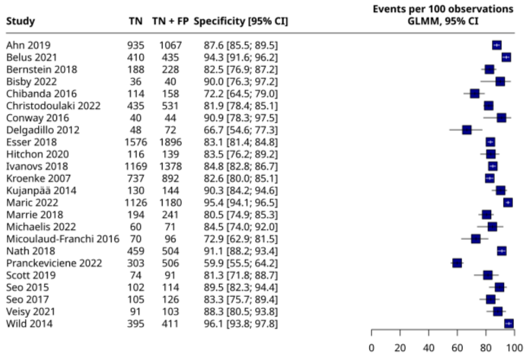


C

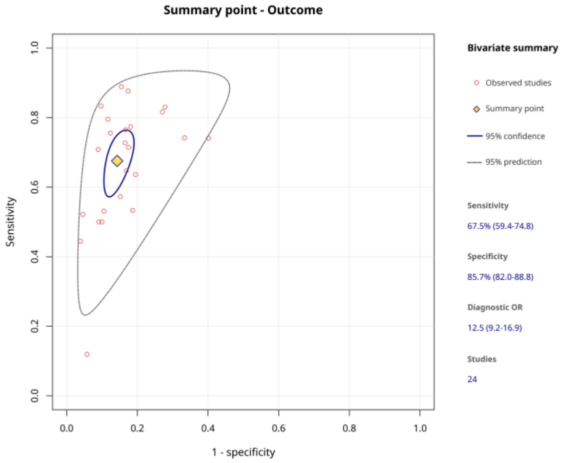


**Figure S7.** Diagnostic test accuracy reproduced in MetaVidence, Akturk et al. (A) Sensitivity; (B) specificity; (C) bivariate summary point.

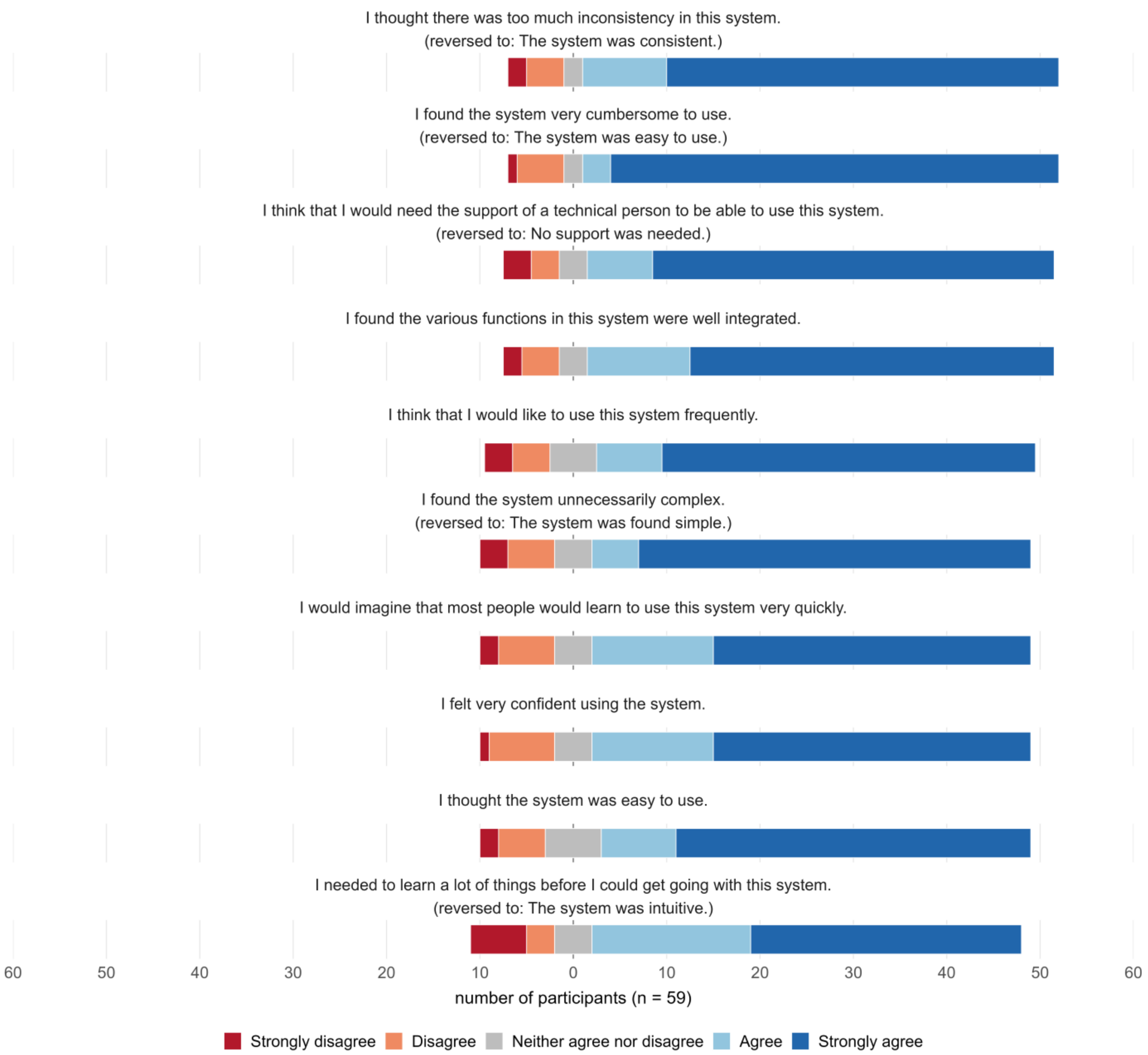


**Figure S8.** Responses to the ten System Usability Scale items (n = 59).

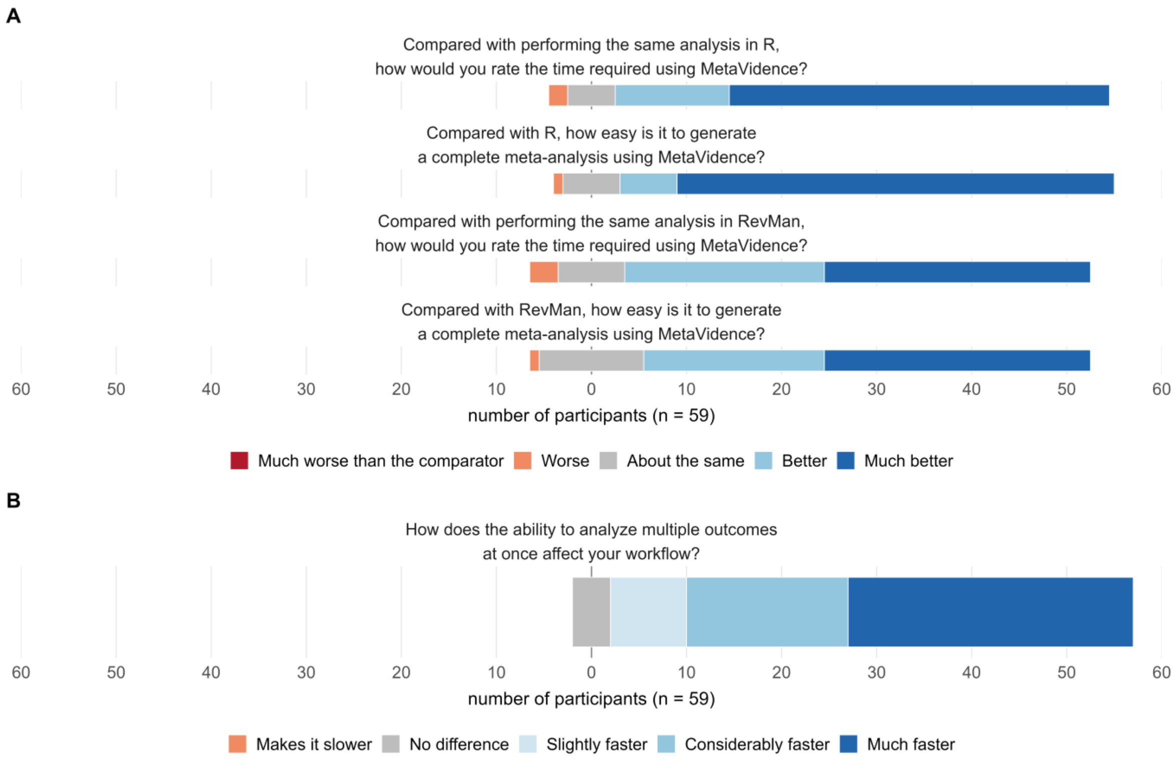


**Figure S9.** (A) Responses to the items comparing MetaVidence with R and with RevMan for the time required and the ease of use; (B) responses to the item on analyzing several outcomes at once (n = 59).

### Appendix S1. Participant instructions and usability questionnaire

#### A. Instructions sent to participants

1. Open https://metavidence.com. The first visit may take up to one minute to load; it opens faster afterwards.

2. Run a meta-analysis of a binary outcome from the first card on the home page, Binary outcome data. You may use the Example tab, which already contains 12 studies, or the spreadsheet attached to the invitation.

3. Then feel free to explore the rest of the application.

4. Finally, complete the questionnaire.

#### B. The questionnaire

MetaVidence is a free, browser-based tool for meta-analysis. This questionnaire takes about 5 minutes.

Participation is voluntary and anonymous: we do not collect your name, e-mail, institution, or any information that could identify you. Responses will be analyzed in aggregate and may be published in a scientific article.

Under Brazilian Resolution CNS 510/2016 (Article 1, sole paragraph, item I), anonymous opinion research is exempt from research ethics committee review.

Questions:

1. How many meta-analyses have you published or conducted

    Open response

**System Usability Scale**

2. I think that I would like to use this system frequently.

    1 Strongly disagree | 2 | 3 | 4 | 5 Strongly agree

3. I found the system unnecessarily complex.

    1 Strongly disagree | 2 | 3 | 4 | 5 Strongly agree

4. I thought the system was easy to use.

    1 Strongly disagree | 2 | 3 | 4 | 5 Strongly agree

5. I think that I would need the support of a technical person to be able to use this system.

    1 Strongly disagree | 2 | 3 | 4 | 5 Strongly agree

6. I found the various functions in this system were well integrated.

    1 Strongly disagree | 2 | 3 | 4 | 5 Strongly agree

7. I thought there was too much inconsistency in this system.

    1 Strongly disagree | 2 | 3 | 4 | 5 Strongly agree

8. I would imagine that most people would learn to use this system very quickly.

    1 Strongly disagree | 2 | 3 | 4 | 5 Strongly agree

9. I found the system very cumbersome (inconvenient) to use.

    1 Strongly disagree | 2 | 3 | 4 | 5 Strongly agree

10. I felt very confident using the system.

    1 Strongly disagree | 2 | 3 | 4 | 5 Strongly agree

11. I needed to learn a lot of things before I could get going with this system.

    1 Strongly disagree | 2 | 3 | 4 | 5 Strongly agree

**Additional items**

12. How does the ability to analyze multiple outcomes at once affect your workflow

    1 Makes it slower | 2 No difference | 3 Slightly faster | 4 Considerably faster | 5 Much faster

13. Compared with performing the same analysis in R, how would you rate the time required using MetaVidence?

    1 Much longer | 2 Longer | 3 About the same | 4 Shorter | 5 Much shorter

14. Compared with R, how easy is it to generate a complete meta-analysis using MetaVidence

    1 Much more difficult | 2 More difficult | 3 About the same | 4 Easier | 5 Much easier

15. Compared with performing the same analysis in RevMan, how would you rate the time required using MetaVidence?

    1 Much longer | 2 Longer | 3 About the same | 4 Shorter | 5 Much shorter

16. Compared with RevMan, how easy is it to generate a complete meta-analysis using MetaVidence

    1 Much more difficult | 2 More difficult | 3 About the same | 4 Easier | 5 Much easier

17. Would you use MetaVidence in a future systematic review or meta-analysis

    1 Not at all | 2 Probably not | 3 Unsure | 4 Probably yes | 5 Definitely yes

18. How likely are you to recommend MetaVidence to another researcher?

    1 Not at all likely | 2 | 3 | 4 | 5 | 6 | 7 | 8 | 9 | 10 Extremely likely

**Scoring**

Items 2 to 11 are the System Usability Scale. Items 2, 4, 6, 8 and 10 are positively worded and items 3, 5, 7, 9 and 11 negatively worded. The score contributed by a positively worded item is its value minus one, and by a negatively worded item is five minus its value; the sum of the ten contributions is multiplied by 2.5 to give a score from 0 to 100.

**Datasets S1 to S12. Study-level input data of the reproduced meta-analyses**

Input data for each of the twelve reproductions in Table 1 of the main text, one table per meta-analysis. Column names match the templates of MetaVidence, so each table can be pasted directly into the application.

**Dataset S1.** Costa Filho et al., 2026.

| Study | events | n |
|---|---|---|
| Divi et al., 2024 | 7 | 20 |
| Ascierto et al., 2023 | 9 | 23 |
| Gross et al., 2023 | 40 | 79 |
| Amatore et al., 2024 | 15 | 26 |

**Dataset S2.** Dacoregio et al., 2026.

| Study | events | n |
|---|---|---|
| PEARL trial | 4 | 50 |
| BELLINI trial b | 2 | 15 |
| NeoTRINNS | 16 | 70 |
| GeparNuevo | 30 | 92 |
| CheckMate 7FL | 92 | 257 |
| IMpassion050 | 107 | 226 |
| neoTAPEL | 20 | 37 |
| IMpassion031 | 93 | 164 |
| KEYNOTE756 | 381 | 634 |
| NeoTRIP | 107 | 138 |
| KEYNOTE522 | 645 | 783 |

**Dataset S3.** Fonseca et al., 2025.

| Study | mean | sd | n |
|---|---|---|---|
| Hanipah et al., 2018 | 42.24 | 21.4 | 11 |
| Sharpton et al., 2019 | 50.75 | 20.8 | 24 |
| Woodford et al., 2015 | 61.3 | 24.1 | 11 |
| Dalal et al., 2004 | 62.7 | 14.9 | 30 |
| Rebibo et al., 2014 | 74.43 | 23.56 | 5 |
| Garcia-Sesma et al., 2019 | 76.3 | 27.55 | 8 |
| Mehdorn et al., 2022 | 79.81 | 33.55 | 27 |

**Dataset S4.** Veeresh et al., 2026.

| Study | mean | sd | n |
|---|---|---|---|
| Brandon W. Smith 2013 | -1.01 | 3.22 | 2400 |
| David J. Mikulis 1992 | -0.06 | 1.78 | 221 |
| Daniel P.M. Barros 2022 | 0.23 | 2.33 | 30 |
| Garland EM 2002 | 0.4 | 2.6 | 46 |
| Braden J. Lawrence 2018 | 0.48 | 2.88 | 10 |
| A.H. Aiken 2012 | 0.7 | 1.9 | 44 |
| Ali Riza Guvercin 2023 | 0.89 | 1.95 | 250 |
| Sheng Yang 2021 | 0.99 | 2.32 | 180 |
| A. J. Barkovich 1986 | 1 | 1.9 | 200 |
| Andreea G. Moore 2016 | 1.5 | 3 | 110 |
| Shamar J. Young 2018 | 1.98 | 2.45 | 200 |
| Kazuhide Furuya 1998 | 2.1 | 2.8 | 9 |
| Ryan K. L. Lee 2015 | 2.2 | 1.8 | 18 |

| Study | mean | sd | n |
|---|---|---|---|
| Aboulezz Ahmed O 1985 | 2.9 | 3.4 | 82 |
| DIPANKAR BISWAS 2019 | 3.01 | 3.42 | 140 |

**Dataset S5.** Defante et al., 2025.

| Study | event.e | n.e | event.c | n.c |
|---|---|---|---|---|
| Iqbal | 28 | 100 | 57 | 100 |
| Morano | 32 | 107 | 58 | 107 |
| Martinez-Galiano | 22 | 70 | 34 | 64 |
| Kettle | 204 | 771 | 338 | 771 |
| Isager-Sally | 156 | 262 | 214 | 261 |
| Mahomed | 129 | 553 | 150 | 524 |
| Almeida | 15 | 48 | 18 | 47 |
| Croce | 40 | 100 | 50 | 102 |
| Kindberg | 65 | 200 | 72 | 200 |
| Sadat-Un-Nisa | 21 | 42 | 38 | 76 |
| Valenzuela | 42 | 223 | 41 | 222 |
| Perveen | 11 | 100 | 10 | 100 |

**Dataset S6.** Katsuyama et al., 2025.

| Study | event.e | n.e | event.c | n.c |
|---|---|---|---|---|
| Xie, 2023 | 0 | 23 | 6 | 33 |
| Shetty, 2015 | 0 | 5 | 1 | 2 |
| Brescia, 2022 | 0 | 37 | 3 | 34 |
| Pfannmueller, 2015 | 1 | 34 | 3 | 22 |
| Baraki, 2013 | 1 | 15 | 2 | 18 |
| Slaughter, 2019 | 48 | 532 | 130 | 962 |
| Gaga, 2013 | 39 | 354 | 75 | 490 |
| Lee, 2020 | 23 | 412 | 21 | 292 |
| Thourani, 2024 | 70 | 1177 | 248 | 3654 |
| Protos, 2018 | 1 | 12 | 1 | 26 |
| Renzulli, 1999 | 1 | 11 | 0 | 10 |

**Dataset S7.** Mamede et al., 2026.

| Study | mean.e | sd.e | n.e | mean.c | sd.c | n.c |
|---|---|---|---|---|---|---|
| Hammond, 2019 | 33.7 | 36.6 | 48 | 57.9 | 79.6 | 48 |
| Hammond, 2018 | 39.01 | 46.71 | 41 | 48.9 | 66.3 | 41 |
| Bakare, 2022 | 20.38 | 21.4 | 22 | 22.44 | 20.92 | 18 |

**Dataset S8.** Ferreira et al., 2023.

| Study | mean.e | sd.e | n.e | mean.c | sd.c | n.c |
|---|---|---|---|---|---|---|
| Granetzky, 2005 | 2 | 4.35 | 20 | 12 | 6.29 | 20 |
| Tanaka, 2002 | 10.8 | 3.4 | 18 | 18.3 | 7.4 | 19 |
| Wu, 2015 | 3.7 | 1.4 | 75 | 9.5 | 4.3 | 89 |
| Dehghan, 2022 | 5.6 | 8.8 | 108 | 6.9 | 10.4 | 99 |
| Marasco, 2013 | 6.32 | 3.46 | 23 | 7.54 | 5.42 | 23 |
| Liu, 2019 | 8.33 | 4.72 | 25 | 9.33 | 3.93 | 25 |

**Dataset S9.** Noronha et al., 2025.

| Study | lnHR | seTE |
|---|---|---|
| Mariani, 2023 | 0.6366 | 0.4521 |
| Hassel, 2023 | 0.7129 | 0.1742 |
| Beasley, 2023 (cohort B) | 1.1378 | 0.777 |

| Study | lnHR | seTE |
| --- | --- | --- |
| Carvajal, 2022 | 1.4702 | 0.2928 |
| Ramelyte, 2024 | 1.5151 | 0.7623 |
| Ny, 2021 | 1.8976 | 0.6827 |
| Rodrigues, 2024 | 2.4078 | 0.7772 |

**Dataset S10.** Lee et al., 2026.

| Study | lnHR | seTE |
| --- | --- | --- |
| Kodama 2024 | -0.1278 | 0.3289 |
| Harada 2023 | -0.0408 | 0.4106 |
| Keum 2024 | 0.239 | 0.1683 |
| Yu 2023 | 0.2546 | 0.0962 |
| Kasi 2021 | 0.27 | 0.3018 |
| Chun 2022 | 0.3436 | 0.2217 |
| Chiu 2022 | 0.4762 | 0.1365 |
| Kawakami 2023 | 0.4824 | 0.3993 |
| Mohring 2023 | 0.6206 | 0.5445 |
| Glassman 2018 | 0.8459 | 0.3213 |
| Okada 2025 | 1.0006 | 0.4575 |
| Yu 2022 | 1.4327 | 0.4228 |

**Dataset S11.** Fomenko et al., 2025.

| Study | TP | FP | FN | TN |
| --- | --- | --- | --- | --- |
| Joling 2012 | 8 | 20 | 2 | 115 |
| Herrero 2003 | 44 | 76 | 6 | 259 |
| Bunevicius 2007 | 110 | 140 | 24 | 229 |
| Lowe 2003 | 43 | 93 | 16 | 198 |
| Bunevicius 2013 | 31 | 116 | 7 | 369 |
| Tiringer 2008 | 57 | 23 | 9 | 129 |
| Fischer 2014 | 18 | 32 | 9 | 135 |
| Sanchez 2014 | 8 | 31 | 3 | 78 |
| Rodriguez-Ramirez 2023 | 357 | 884 | 97 | 1365 |
| Yamamoto-Furusho 2021 | 14 | 14 | 7 | 69 |
| Bernstein 2018 | 31 | 58 | 11 | 141 |
| Preljevic 2012 | 11 | 16 | 7 | 73 |
| Vasquez 2013 | 12 | 12 | 6 | 86 |
| Castro 2006 | 39 | 28 | 4 | 20 |
| Reme 2014 | 40 | 81 | 29 | 395 |
| Gandy 2015 | 27 | 26 | 17 | 77 |
| Sanchez-Gistau 2012 | 60 | 132 | 18 | 195 |
| Al-Asmi 2012 | 56 | 7 | 10 | 73 |
| Marrie 2018 | 28 | 58 | 15 | 151 |
| Leentjens 2011 | 90 | 96 | 28 | 127 |
| McWhirter 2021 | 9 | 8 | 2 | 29 |
| Sagen 2009 | 12 | 8 | 12 | 69 |
| O'Rourke 1998 | 9 | 18 | 4 | 74 |
| McKenzie 2018 | 24 | 18 | 11 | 85 |
| Whelan-Goodinson 2009 | 27 | 20 | 9 | 44 |
| Patel 2011 | 5 | 29 | 2 | 59 |
| Nakash 2020 | 4 | 25 | 1 | 56 |
| Patel 2010 | 5 | 16 | 1 | 56 |
| Van Beek 2022 | 6 | 102 | 5 | 333 |
| Singer 2008 | 7 | 53 | 1 | 210 |
| Esser 2018 | 137 | 475 | 112 | 1418 |
| Clover 2020 | 23 | 19 | 14 | 76 |

| Study | TP | FP | FN | TN |
|---|---|---|---|---|
| Requena 2009 | 15 | 50 | 6 | 163 |
| Singer 2018 | 15 | 175 | 3 | 590 |
| Singer 2009 | 43 | 215 | 8 | 325 |
| Colomo 2021 | 20 | 8 | 5 | 19 |
| Cheung 2012 | 5 | 4 | 9 | 37 |
| Baker 2018 | 15 | 28 | 9 | 168 |
| Phan 2016 | 10 | 8 | 4 | 34 |
| Soyseth 2016 | 12 | 5 | 19 | 58 |
| Law 2014 | 27 | 29 | 2 | 43 |
| Wong 2015 | 27 | 16 | 1 | 71 |
| Hitchon 2020 | 23 | 38 | 5 | 83 |
| Lok 2022 | 23 | 45 | 3 | 129 |
| Chan 2017a | 19 | 19 | 6 | 116 |
| Kwan 2019 | 15 | 7 | 8 | 20 |

**Dataset S12.** Akturk et al., 2025.

| Study | TP | FP | FN | TN |
|---|---|---|---|---|
| Maric 2022 | 12 | 54 | 11 | 1126 |
| Christodoulaki 2022 | 41 | 96 | 12 | 435 |
| Pranckeviciene 2022 | 40 | 203 | 14 | 303 |
| Wild 2014 | 12 | 16 | 15 | 395 |
| Ahn 2019 | 68 | 132 | 22 | 935 |
| Belus 2021 | 8 | 25 | 59 | 410 |
| Ivanovs 2018 | 51 | 209 | 38 | 1169 |
| Chibanda 2016 | 88 | 44 | 18 | 114 |
| Kroenke 2007 | 64 | 155 | 9 | 737 |
| Kujanpaa 2014 | 5 | 14 | 1 | 130 |
| Scott 2019 | 8 | 17 | 7 | 74 |
| Micoulaud-Franchi 2016 | 40 | 26 | 9 | 70 |
| Michaelis 2022 | 8 | 11 | 1 | 60 |
| Bisby 2022 | 11 | 4 | 11 | 36 |
| Seo 2015 | 17 | 12 | 15 | 102 |
| Veisy 2021 | 66 | 12 | 17 | 91 |
| Seo 2017 | 26 | 21 | 8 | 105 |
| Marrie 2018 | 7 | 47 | 4 | 194 |
| Esser 2018 | 22 | 320 | 12 | 1576 |
| Conway 2016 | 2 | 4 | 2 | 40 |
| Nath 2018 | 17 | 45 | 7 | 459 |
| Delgadillo 2012 | 23 | 24 | 8 | 48 |
| Bernstein 2018 | 10 | 40 | 4 | 188 |
| Hitchon 2020 | 8 | 23 | 3 | 116 |
